# Adult Vaccination History and Plasma Biomarker Signatures in the Asymptomatic PREVENT-AD Cohort

**DOI:** 10.64898/2026.08.24.26361238

**Authors:** Sophie Charland, Mélissa Savard, Isabel Sarty, Christine Dery, Sylvia Villeneuve, Cynthia Picard, Judes Poirier

## Abstract

**Background:** Epidemiological studies increasingly associate several adult vaccinations with lower risk of Alzheimer’s disease (AD) and dementia, but the biological mechanisms underlying these observations remain unclear. We investigated whether adult vaccination history is associated with AD-related and immune-related biomarker profiles in cognitively unimpaired individuals at increased familial risk for AD.

**Methods:** This exploratory study included 192 participants from the PREVENT-AD cohort. Adult vaccination and infection histories were collected using a structured questionnaire and examined in relation to plasma and cerebrospinal fluid (CSF) biomarkers. Adjusted regression models evaluated individual vaccine exposures, vaccination-profile breadth, broader proteomic signatures, CSF biomarkers, viral-history interactions, and psychological symptoms, with false-discovery-rate (FDR) correction applied for multiple testing.

**Results:** Influenza, herpes zoster, pneumococcal, and Td/Tdap vaccination were not associated with FDR-significant differences in the principal plasma amyloid and tau biomarker panel. Hepatitis B vaccination was associated with lower plasma total tau/MAPT, p-tau181, and p-tau231 after correction within the AD biomarker panel, although these findings may reflect residual behavioral or healthcare-related confounding. Greater vaccination-profile breadth was associated with higher plasma NPTX1 (β = 0.261, p = 0.007, q = 0.049), whereas its nominal association with a lower Aβ42/Aβ40 ratio did not survive FDR correction. Nominal herpes zoster associations with lower CSF Aβ42 and pTau were similarly attenuated after correction. No robust FDR-significant associations emerged from viral-history interaction or psychological symptom analyses.

**Conclusions:** Adult vaccination history was not associated with a broad plasma amyloid or tau signature in this asymptomatic, familial-risk cohort. However, the association between broader vaccination exposure and higher NPTX1 suggests a potentially distinct synaptic-related signal, while the hepatitis B findings identify additional hypothesis-generating tau-related associations. Longitudinal studies incorporating vaccine timing, infection burden, and repeated biomarker measurements are needed to determine whether vaccination influences biological pathways relevant to AD resilience.

## INTRODUCTION

Alzheimer’s disease (AD) is biologically defined by the progressive accumulation of amyloid-β and tau pathology, yet the processes that determine when these lesions become clinically consequential remain incompletely understood. Over the last three decades, a substantial body of work has suggested that infectious exposures, pathogen reactivation, and host immune responses may modify the risk, timing, or trajectory of AD and related dementias. The prevailing view in the field now acknowledges that Alzheimer’s disease isn’t caused by just one virus or bacterium. Rather, it is viewed as a complicated, multifaceted condition involving many interacting factors over long durations. These factors comprise dormant infections, recurring systemic inflammation, age-related immune weakening, APOE gene variations, compromised blood vessel performance, a weakened blood-brain barrier, and pre-existing amyloid and tau protein accumulation. The combination of these elements impacts the brain’s resilience and when the disease’s symptoms manifest. Consistent with the heterogeneity of late-onset AD, this refined framework matches observations where infection-related links are found across all types of dementia, mixed dementia syndromes, and cognitive decline, not solely in pathologically pure AD. (Ukraintseva et al., 2024)

Among infectious candidates, herpesviruses have received particular attention. Herpes simplex virus type 1 (HSV-1) has been detected in a high proportion of older human brains, and early studies proposed that the combination of HSV-1 brain infection and APOE ε4 carriage may increase AD risk(Itzhaki, Wozniak, Appelt, & Balin, 2004). Subsequent work has linked HSV-1 reactivation, indexed by serological markers, to increased future AD risk, while mechanistic studies have shown that HSV-1 can engage several core AD-relevant processes, including amyloid-β production, tau phosphorylation, oxidative stress, metabolic dysfunction, and glial activation(Liu, Johnston, Jarousse, Fletcher, & Iqbal, 2025). Varicella-zoster virus (VZV), another lifelong latent herpesvirus, has also become highly relevant because zoster reactivation is common in older adults and has been associated with later cognitive outcomes(Cairns, Itzhaki, & Kaplan, 2022; Polisky et al., 2025). Together, these observations support the hypothesis that repeated viral reactivation may act less as a direct infectious cause of AD than as a chronic neuroimmune stressor capable of lowering the threshold for clinical disease in biologically susceptible individuals.

The study of bacteria has progressed concurrently. Researchers found Chlamydia pneumoniae in Alzheimer’s disease brain tissue, and newer studies connect it to inflammation, amyloid and tau issues, APOE ε4, and memory loss(Gaire et al., 2026; Subedi, Gaire, Koronyo, Koronyo-Hamaoui, & Crother, 2024). The oral-brain axis has also emerged as a major area of interest, particularly through periodontitis and Porphyromonas gingivalis, where epidemiological studies, brain-tissue analyses, and experimental models converge on mechanisms involving bacterial products, gingipains, complement activation, microglial stimulation, amyloidogenic processing, and impaired clearance. Other organisms, including Helicobacter pylori and Borrelia species, have been associated with dementia or AD in selected studies, but the evidence remains more heterogeneous(Ahuja, Shaban, Chawla, & Parmar, 2026; Kelsey et al., 2025; Xie et al., 2023). Overall, the infectious-exposure literature is strongest when interpreted as evidence that chronic or recurrent inflammatory burden may amplify AD pathophysiology, rather than as proof of a simple pathogen-specific etiology.

A major conceptual advance in this field has been the reinterpretation of amyloid-β, and possibly tau, as components of innate antimicrobial defense(Eimer et al., 2026; Kumar et al., 2016). Experimental studies showing that amyloid-β can protect against microbial infection provided a mechanistic bridge between infection biology and one of the defining pathological features of AD. In this model, amyloid and tau responses may initially represent adaptive host-defense mechanisms, but repeated or chronic induction in the aging brain could become maladaptive, leading to persistent glial activation, complement engagement, vascular injury, synaptic dysfunction, and impaired clearance. This framework is particularly attractive because it links infectious exposures to established AD biology without requiring that pathogens be continuously present or uniquely causal at the time of clinical diagnosis.

The strongest recent epidemiological development has been the observation that several adult vaccinations are associated with reduced dementia or AD risk. Influenza vaccination has repeatedly been associated with lower incident AD or dementia risk in claims-based and meta-analytic studies, with some evidence for dose-response effects and potentially stronger associations in higher-risk populations(Bukhbinder et al., 2022; Bukhbinder et al., 2026; Yang & Jiang, 2026). Pneumococcal vaccination has also been linked to lower AD risk in older adults, with possible genotype-dependent effects(Huo & Finkelstein, 2024; Ukraintseva et al., 2023). Tdap/Td vaccination has been associated with reduced all-cause dementia or AD risk in multiple cohorts.(Harris et al., 2023; Scherrer et al., 2021) Although these findings are susceptible to healthy-vaccinee bias, healthcare-engagement bias, and residual confounding, the consistency across unrelated vaccine classes suggests that vaccination may act through more than simple prevention of one specific pathogen. Instead, vaccination could reduce the cumulative infection burden, attenuate systemic inflammatory insults, modulate innate immune set-points, or enhance immune surveillance in ways that are relevant to aging-brain vulnerability(Ukraintseva et al., 2024).

The most interesting vaccine evidence currently comes from herpes zoster vaccination. A natural experiment in Wales using a birth-date eligibility cutoff for the live zoster vaccine found an approximately 20% relative reduction in new dementia diagnoses over seven years, with stronger effects in women(Eyting et al., 2025). Because this design is less vulnerable to healthy-vaccinee bias than conventional observational comparisons, it represents unusually persuasive evidence that zoster vaccination may modify dementia risk. More recently, large comparative cohort studies have suggested that recombinant zoster vaccine, including Shingrix, is associated with longer dementia-free survival and lower dementia risk than live zoster vaccine or comparator vaccines(Duan et al., 2026; Rayens et al., 2026; Taquet, Dercon, Todd, & Harrison, 2024). The evidence base supports live zoster vaccination as moderate-to-strong evidence for reduced dementia incidence, while recombinant zoster vaccine is supported by moderate evidence because the findings are consistent across large datasets but remain observational.

The COVID-19 pandemic further reinforced the relevance of infection-related neuroimmune stress to late-life cognition. Large electronic-health-record and community cohorts have reported persistent cognitive symptoms, accelerated decline after severe or hospitalized COVID-19, and increased short-term dementia diagnoses following SARS-CoV-2 infection(Demmer et al., 2025; Wang et al., 2022). CSF and plasma studies indicate that COVID-19 can induce inflammatory, neuroaxonal, vascular, and AD-like biomarker changes, including reduced plasma Aβ42:Aβ40 ratio and, in vulnerable participants, higher p-tau181(Duff et al., 2025). These findings do not establish that SARS-CoV-2 causes de novo AD, particularly given the long prodromal phase of the disease, but they strongly support the concept that systemic viral illness can accelerate or unmask pre-existing neurodegenerative processes through inflammation, endothelial injury, blood-brain barrier dysfunction, glial priming, hypoxia, and critical-illness effects.

Despite this expanding literature, major gaps remain. Most vaccine studies rely on clinical diagnostic codes rather than biomarker-confirmed AD. Few studies have access to longitudinal cerebrospinal fluid, plasma, neuroimaging, and cognitive measures obtained before and after vaccination. Even fewer can examine vaccination in asymptomatic individuals at increased familial risk, where effects on preclinical AD biology may be detectable before diagnostic endpoints emerge. This gap is important because the negative valacyclovir trial in early symptomatic AD suggests that late antimicrobial intervention is unlikely to reverse established self-propagating amyloid, tau, and neurodegenerative cascades, whereas immune modulation or infection prevention may be more relevant during the long preclinical phase.

The PREVENT-AD cohort offers a unique opportunity to address this mechanistic gap. PREVENT-AD is a longitudinal cohort of cognitively unimpaired older adults enriched for parental history of AD, with repeated clinical assessments and extensive biofluid sampling over many years(Villeneuve et al., 2025). Building on the infection-vaccination literature, we examined whether adult vaccination history, including shingles, COVID-19, influenza, and Tdap vaccination, is associated with biochemical profiles in cerebrospinal fluid and plasma. Furthermore, travel-related vaccinations have also been inadequately studied in this area. Rather than asking whether vaccination merely predicts future diagnostic status, we investigated whether vaccination is linked to AD pathological biomarkers and immune-related molecular signatures in bio-fluids from asymptomatic individuals at increased risk. This approach is well-suited to test whether vaccination-associated protection may involve modulation of inflammatory pathways, innate immune tone, neurodegeneration-related proteins, or early AD biomarker trajectories before the onset of cognitive impairment.

Here, we present a pilot analysis of vaccination exposure in PREVENT-AD and its relationship to cross-sectional bio-fluid markers of AD pathophysiology and inflammation. We hypothesized that vaccination would be associated with a more favorable molecular profile, potentially reflecting reduced infection burden, altered systemic immune tone, or vaccine-induced remodeling of innate and adaptive immune responses. By integrating vaccination history with CSF and plasma biomarkers in a deeply phenotyped asymptomatic cohort, this study aims to move the field beyond epidemiological associations and toward a mechanistic understanding of how adult vaccination may influence biological pathways relevant to AD onset and resilience.

## Materials and Methods

### Study design and participants

This exploratory analysis was conducted in the PREVENT-AD cohort, a longitudinal, single-site study of cognitively unimpaired older adults enriched for familial risk of sporadic Alzheimer’s disease. Participants were recruited from the greater Montreal area and were eligible if they had a first-degree family history of sporadic Alzheimer-like dementia or multiple affected siblings and were cognitively normal at study entry. The present analysis used a de-identified participant-level analytic file containing vaccination history, demographic variables, APOE4 status where available, and plasma/CSF biomarker measurements. A subset of 192 participants has agreed to respond to the survey.

### Vaccination and viral infection history

Vaccination and infection history were obtained using a structured REDCap web-questionnaire in the summer of 2026. The survey was designed to capture adult vaccination history for seasonal influenza, herpes zoster/shingles, pneumococcal vaccination, tetanus-diphtheria-pertussis/tetanus-diphtheria (Td/Tdap) vaccination, COVID-19, hepatitis A, hepatitis B, RSV/VRS and other adult vaccines. For zoster vaccination, the questionnaire distinguished Zostavax, Shingrix, undetermined zoster vaccine type, and no vaccination. For pneumococcal vaccination, Pneumovax, Prevnar 13, Prevnar 20 and unknown type were captured. The same form also queried about known viral infection history, including varicella, shingles, recurrent herpes simplex virus type 1 (HSV-1), genital herpes/possible HSV-2, EBV/mononucleosis, CMV, serious influenza, viral pneumonia, COVID-19, viral meningitis/encephalitis/CNS infection, hepatitis A/C, hepatitis B, severe viral infection requiring hospitalization and perceived persistent cognitive worsening after major viral infection.

The primary vaccine exposures examined were influenza, zoster/shingles, pneumococcal, Td/Tdap, hepatitis A and hepatitis B vaccination. COVID-19 vaccination was summarized descriptively but was not modeled because nearly all participants were vaccinated at one moment or another (190 of 192), leaving only two unvaccinated participants. RSV/VRS vaccination was also summarized descriptively but was not prioritized for regression modeling because it was uncommon in this dataset. However, it was included in the additional exploratory individual-vaccine analyses.

### Biomarker outcomes

AD-related biomarker outcomes included CSF ELISA measures of Aβ42, total tau and phosphorylated tau (Innotest, Fujirebio, Sweden), together with plasma NULISA measures of Aβ38, Aβ40, Aβ42, p-tau181, p-tau217, p-tau231, BD p-tau181 and total tau/MAPT (Alamar, USA). Because NULISA values are expressed on a log2-like scale in the analytic file, a plasma Aβ42:Aβ40 index was calculated as the log2 difference between plasma Aβ42 and plasma Aβ40. The full plasma NULISA panel included 115 measured analytes spanning amyloid/tau biology, neurodegeneration, synaptic proteins, cytokines, chemokines, vascular markers and glial/inflammatory proteins. The Olink panel included 12 candidate markers recently identified as modulators of tau physiology that impact synaptic integrity: CNTN5, RGMB, LAYN, CD200, EPHB6, NTRK2, SCARB2, ADAM23, SIRT2, KITLG, PDGFRA and CDH6 (resubmitted, Nature Molecular Psychiatry 2026).

A focused exploratory plasma biomarker panel was additionally examined and included Aß40, Aß42, Aß42/Aß40 ratio, pTau217, pTau181, GFAP, and NPTX1. Exploratory CSF replication analyses were restricted to the available CSF Aß42 and phosphorylated tau measurement.

### Statistical analysis

Descriptive statistics were calculated for demographic variables and vaccination exposures. Associations between individual vaccination exposures and biomarker levels were tested using separate linear regression models for each biomarker. The primary model was: standardized biomarker level = vaccination status + age at baseline + sex + education years + APOE4 carrier status. Additional exploratory analyses of individual vaccine exposures, vaccination profile breadth, CSF replication, viral history interaction, and psychological symptoms were conducted in Jamovi 2025 and adjusted for age, sex, and APOE4 status. Education was not included as a covariate in these exploratory models. Assumptions of homogeneity of variance were assessed using Levene’s test. When Levene’s test indicated heterogeneity of variance (p<0.05), robustness of analysis was performed using heteroskedasticity-robust standard errors.

Biomarker outcomes were standardized before regression so that the vaccination coefficient represents the adjusted difference in standard deviation units between vaccinated and unvaccinated participants. Unadjusted vaccinated and unvaccinated biomarker means were also calculated for interpretability. Complete-case analysis was used; therefore, the sample size varies by vaccine, biomarker, and covariate availability. Models were not interpreted when the effective vaccinated or unvaccinated group contained fewer than approximately 10 participants, except for the small Olink exploratory subset where results are reported as descriptive/exploratory.

Multiple-testing correction was performed using the Benjamini-Hochberg false-discovery-rate (FDR) procedure within each vaccine: the AD biomarker panel, the full NULISA panel, and the Olink panel. For the additional exploratory analyses, FDR correction was applied across all tests within each predefined analysis family, regardless of the nominal significance. These families included individual vaccine– plasma biomarker associations, vaccination-profile breadth analyses, CSF replication analyses, viral-history interaction analyses, and psychological symptom analyses.

All analyses are exploratory and should be interpreted as hypothesis-generating. The analysis is not designed to infer causality because vaccination was self-reported, vaccine timing relative to biomarker collection was not modeled, and co-vaccination/healthy-vaccinee effects are likely.

## Results

### Analytic sample and vaccination exposure distribution

Cross-sectional analyses were performed on a total of 192 participant records (Table 1). The mean baseline age was 62.5 years (SD 4.4), 126 participants were female and 64 were male among those with available sex information, and mean education was 15.7 years (SD 3.1). APOE4 status was available for 148 participants, of whom 59 were APOE4 carriers. At the available baseline/proximal visit, 171 participants were classified as cognitively normal and 19 as MCI in the analytic file.

**Table 1.**
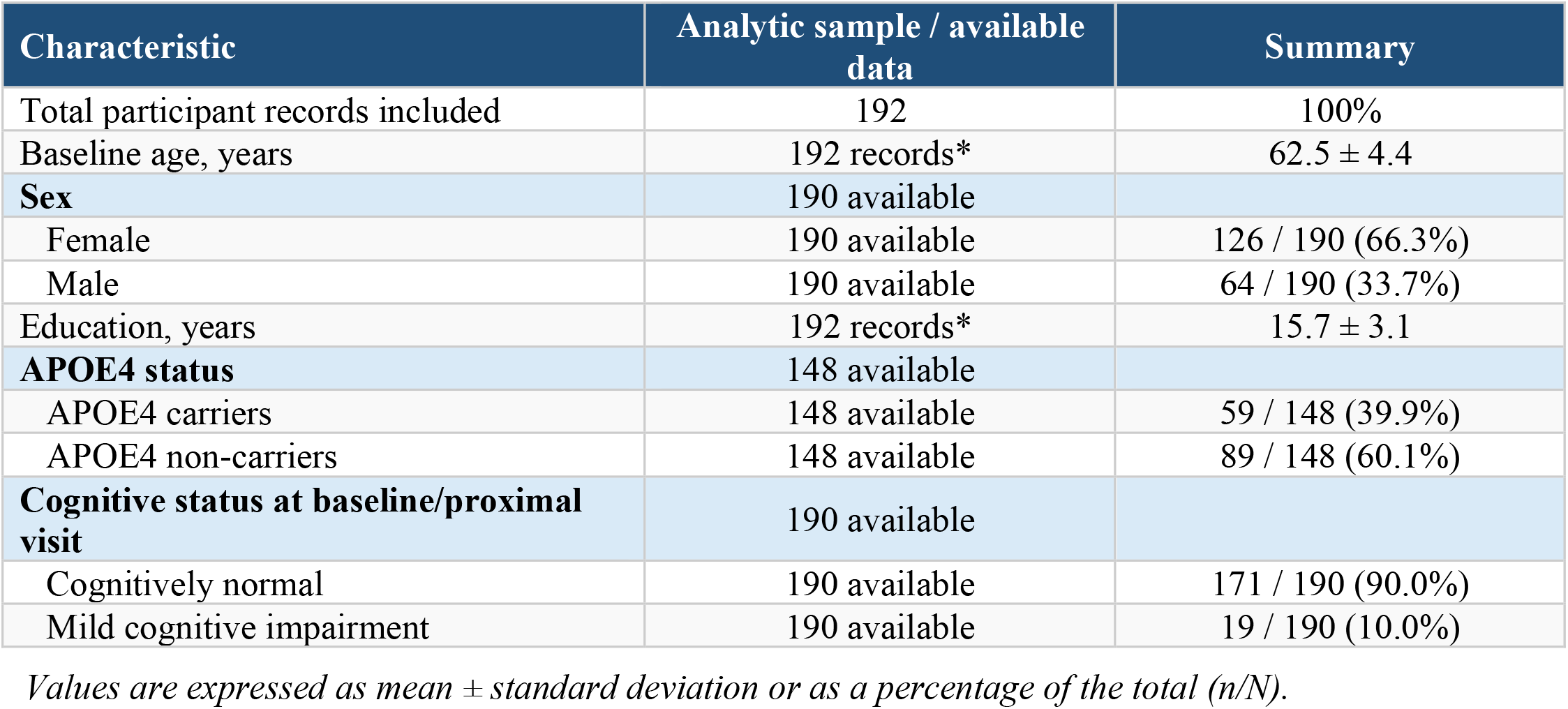
Demographic and clinical characteristics.

| Characteristic | Analytic sample / available data | Summary |
| --- | --- | --- |
| Total participant records included | 192 | 100% |
| Baseline age, years | 192 records* | 62.5 ± 4.4 |
| **Sex** | 190 available |  |
| Female | 190 available | 126 / 190 (66.3%) |
| Male | 190 available | 64 / 190 (33.7%) |
| Education, years | 192 records* | 15.7 ± 3.1 |
| **APOE4 status** | 148 available |  |
| APOE4 carriers | 148 available | 59 / 148 (39.9%) |
| APOE4 non-carriers | 148 available | 89 / 148 (60.1%) |
| **Cognitive status at baseline/proximal visit** | 190 available |  |
| Cognitively normal | 190 available | 171 / 190 (90.0%) |
| Mild cognitive impairment | 190 available | 19 / 190 (10.0%) |

*Values are expressed as mean ± standard deviation or as a percentage of the total (n/N).*

**Table 2.** Vaccination exposure distribution.

| Vaccine | Vaccinated (n) | Unvaccinated (n) | Missing/unknown (n) | Vaccinated % among known |
| --- | --- | --- | --- | --- |
| Influenza | 156 | 36 | 0 | 81.2% |
| Zoster/shingles | 150 | 41 | 1 | 78.5% |
| Pneumococcal | 128 | 56 | 8 | 69.6% |
| Td/Tdap | 135 | 28 | 29 | 82.8% |
| COVID-19 | 190 | 2 | 0 | 99.0% |
| Hepatitis A | 87 | 71 | 34 | 55.1% |
| Hepatitis B | 86 | 70 | 36 | 55.1% |
| RSV/VRS | 19 | 145 | 28 | 11.6% |

**Table 3.** AD biomarker associations reaching nominal P < 0.05 in adjusted models.

| Vaccine | AD biomarker | N | Vaccinated<br>Unvaccinated<br>(n) | Mean<br>vacc | Mean<br>unvacc | Adj<br>beta_std | Nominal<br>P value | BH q* |
| --- | --- | --- | --- | --- | --- | --- | --- | --- |
| Hepatitis B | total<br>tau/MAPT | 111 | 55/56 | 11.595 | 11.915 | -0.573 | 0.0029 | 0.0349 |
| Hepatitis A | p-tau231 | 111 | 56/55 | 12.386 | 12.713 | -0.518 | 0.0050 | 0.0525 |
| Hepatitis B | p-tau181 | 111 | 55/56 | 12.270 | 12.559 | -0.463 | 0.0102 | 0.0466 |
| Hepatitis B | p-tau231 | 111 | 55/56 | 12.384 | 12.687 | -0.465 | 0.0116 | 0.0466 |
| Hepatitis A | total tau/MAPT | 111 | 56/55 | 11.643 | 11.922 | -0.483 | 0.0129 | 0.0525 |
| Hepatitis A | p-tau181 | 111 | 56/55 | 12.290 | 12.578 | -0.452 | 0.0131 | 0.0525 |
| Influenza | Aβ40 | 139 | 118/21 | 11.311 | 11.041 | 0.496 | 0.0401 | 0.3909 |
| Hepatitis B | Aβ42 | 111 | 55/56 | 12.588 | 12.902 | -0.392 | 0.0459 | 0.1377 |
| Hepatitis A | Aβ42 | 111 | 56/55 | 12.582 | 12.900 | -0.388 | 0.0474 | 0.1422 |

*Note. Only nominally significant associations are shown. BH $q^*$ values (after FDR corrections) were corrected within the AD biomarker panel for each vaccine. Positive beta\_std indicates higher biomarker level in vaccinated participants; negative beta\_std indicates lower biomarker level.*

**Table 4.** Plasma NULISA associations reaching nominal P < 0.05.

| Vaccine | NULISA biomarker | N | Vacc/Unvacc (n) | Adj beta_std | Nominal P value* | BH q* |
| --- | --- | --- | --- | --- | --- | --- |
| Pneumococcal | CALB2 | 133 | 102/31 | 0.602 | 0.0025 | 0.2833 |
| Hepatitis B | MAPT | 111 | 55/56 | -0.573 | 0.0029 | 0.3340 |
| Hepatitis A | pTau 231 | 111 | 56/55 | -0.518 | 0.0050 | 0.2154 |
| Hepatitis A | NPY | 111 | 56/55 | -0.519 | 0.0072 | 0.2154 |
| Hepatitis A | IL6R | 111 | 56/55 | -0.526 | 0.0074 | 0.2154 |
| Hepatitis A | POSTN | 111 | 56/55 | -0.499 | 0.0088 | 0.2154 |
| Influenza | VEGFA | 139 | 118/21 | 0.628 | 0.0091 | 0.6032 |
| Hepatitis B | pTau 181 | 111 | 55/56 | -0.463 | 0.0102 | 0.3395 |
| Zoster/shingles | CCL11 | 138 | 110/28 | -0.547 | 0.0108 | 0.8534 |
| Hepatitis B | pTau 231 | 111 | 55/56 | -0.465 | 0.0116 | 0.3395 |
| Hepatitis A | PARK7 | 111 | 56/55 | 0.492 | 0.0120 | 0.2154 |
| Hepatitis A | MAPT | 111 | 56/55 | -0.483 | 0.0129 | 0.2154 |
| Hepatitis A | pTau 181 | 111 | 56/55 | -0.452 | 0.0131 | 0.2154 |
| Hepatitis B | NPY | 111 | 55/56 | -0.474 | 0.0138 | 0.3395 |
| Hepatitis B | PARK7 | 111 | 55/56 | 0.479 | 0.0148 | 0.3395 |
| Influenza | CALB2 | 139 | 118/21 | 0.560 | 0.0148 | 0.6032 |
| Influenza | CHIT1 | 134 | 115/19 | 0.610 | 0.0157 | 0.6032 |
| Pneumococcal | CD63 | 133 | 102/31 | -0.484 | 0.0167 | 0.6061 |
| Pneumococcal | BACE1 | 133 | 102/31 | -0.490 | 0.0183 | 0.6061 |
| Pneumococcal | DDC | 133 | 102/31 | -0.476 | 0.0227 | 0.6061 |
| Influenza | CXCL1 | 139 | 118/21 | 0.543 | 0.0240 | 0.6887 |
| Pneumococcal | ENO2 | 133 | 102/31 | -0.465 | 0.0264 | 0.6061 |
| Zoster/shingles | FABP3 | 138 | 110/28 | -0.452 | 0.0306 | 0.8534 |
| Hepatitis B | POSTN | 111 | 55/56 | -0.413 | 0.0309 | 0.5915 |
| Hepatitis A | VEGFD | 111 | 56/55 | 0.420 | 0.0318 | 0.4572 |
| Pneumococcal | CCL3 | 133 | 102/31 | 0.433 | 0.0375 | 0.6512 |
| Zoster/shingles | SLIT2 | 138 | 110/28 | -0.447 | 0.0399 | 0.8534 |
| Influenza | A 40 | 139 | 118/21 | 0.496 | 0.0401 | 0.7467 |
| Hepatitis B | A 42 | 111 | 55/56 | -0.392 | 0.0459 | 0.7540 |
| Hepatitis A | A 42 | 111 | 56/55 | -0.388 | 0.0474 | 0.6058 |

*No NULISA association survived BH\* correction across the full 115-analyte NULISA panel. Table is intended to prioritize replication candidates.*

**Table 5.** Olink biomarker associations for zoster/shingles vaccination.

| Olink biomarker | N | Vacc/Unvacc n | Mean zoster-vacc | Mean unvacc | Adj beta_std | P value | BH q |
| --- | --- | --- | --- | --- | --- | --- | --- |
| NTRK2 | 41 | 32/9 | 3.765 | 3.537 | 0.585 | 0.1384 | 0.4117 |
| CD200 | 41 | 32/9 | 4.383 | 4.165 | 0.516 | 0.2128 | 0.4117 |
| CDH6 | 41 | 32/9 | 0.650 | 0.390 | 0.502 | 0.2145 | 0.4117 |
| RGMB | 41 | 32/9 | 5.360 | 5.102 | 0.492 | 0.2293 | 0.4117 |
| KITLG | 41 | 32/9 | -2.152 | -2.447 | 0.485 | 0.2308 | 0.4117 |
| PDGFRA | 41 | 32/9 | 1.963 | 1.736 | 0.503 | 0.2418 | 0.4117 |
| SCARB2 | 41 | 32/9 | 0.831 | 0.691 | 0.476 | 0.2557 | 0.4117 |
| ADAM23 | 41 | 32/9 | -0.776 | -0.919 | 0.472 | 0.2745 | 0.4117 |
| CNTN5 | 41 | 32/9 | 3.426 | 3.220 | 0.381 | 0.3473 | 0.4631 |
| EPHB6 | 41 | 32/9 | 1.035 | 0.817 | 0.351 | 0.4040 | 0.4849 |
| LAYN | 41 | 32/9 | 4.054 | 3.890 | 0.307 | 0.4630 | 0.5051 |
| SIRT2 | 41 | 32/9 | -0.645 | -0.739 | 0.214 | 0.6138 | 0.6138 |

*Note. The Olink subset was small; none of the zoster/shingles associations reached nominal $P < 0.05$ or FDR significance.*

Vaccination rates were high for influenza, zoster/shingles, Td/Tdap and COVID-19. Zoster vaccination was reported by 150 of 191 participants with known zoster-vaccine status; among categorical responses, 124 reported Shingrix, 5 Zostavax and 21 unknown zoster-vaccine type. COVID-19 vaccination was nearly universal, preventing meaningful vaccinated-versus-unvaccinated regression analysis. Vaccination against Hepatitis A and B, though less frequent, frequently occurred together, aligning with patterns related to travel, occupation, or healthcare access (supplemental table 1).

### Associations with AD amyloid and tau biomarkers

No robust FDR-significant association was observed for influenza, zoster/shingles, pneumococcal or Td/Tdap vaccination in the AD biomarker panel after adjustment for age, sex, education and APOE4 status. The most consistent signal was observed for hepatitis vaccination, especially hepatitis B, which was associated with lower plasma tau-related biomarkers. Hepatitis B vaccination was associated with lower plasma total tau/MAPT, lower plasma p-tau181 and lower plasma p-tau231 after correction within the AD biomarker panel. A similar hepatitis A pattern was observed at nominal significance but did not quite meet FDR significance in the AD panel. These hepatitis-vaccine associations should be interpreted cautiously because hepatitis A and B vaccination were highly correlated and may index unmeasured health-behavior, travel, occupational or healthcare-engagement factors rather than a pathogen-specific AD mechanism. Standardized effect estimates for the individual vaccination-plasma biomarker associations are summarized in Figure 1, with full regression results provided in Supplementary Table S2

**Figure 1.**
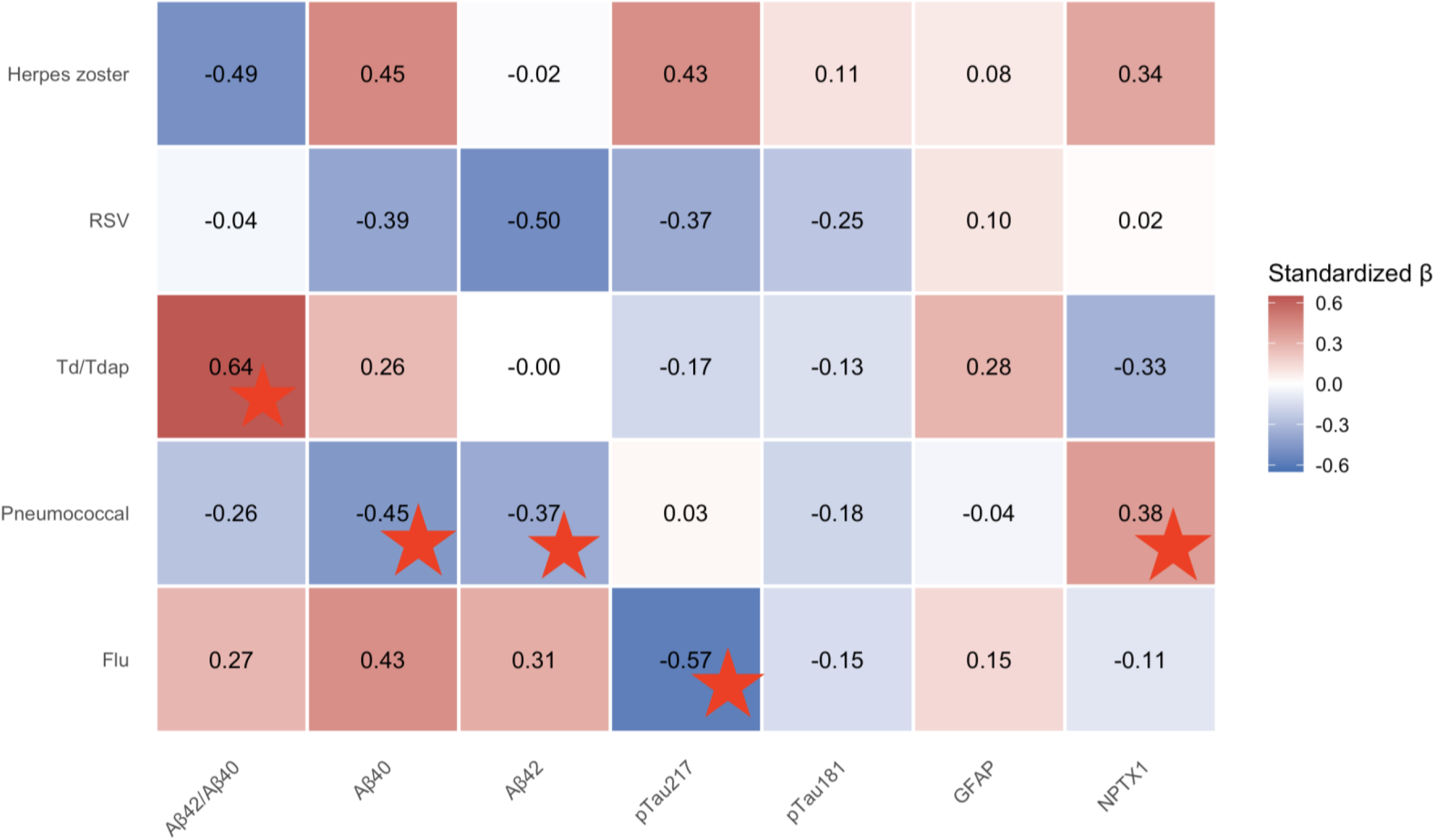
Adjusted associations between individual adult vaccination exposures and plasma biomarkers. Cells represent standardized β coefficients from regression models adjusted for age, sex, and APOE4 carrier status. Positive values indicate higher biomarker levels in vaccinated participants, whereas negative values indicate lower levels. Stars indicate nominal significance before FDR correction.

### Associations with vaccination profile breadth

Vaccination-profile breadth was defined as the number of distinct vaccine types received across the life course rather than the number of vaccine doses. A secondary analysis was conducted to evaluate associations between vaccination-profile breadth and Alzheimer’s disease-related plasma biomarkers (Supplementary Table S3). Seven adjusted regression models were performed. Two nominal associations were observed. Greater vaccination-profile breadth was associated with a lower plasma Aβ42/Aβ40 ratio (β = −0.211, p = 0.034), although this association did not remain significant after FDR correction (q = 0.119). Greater vaccination-profile breadth was also associated with higher plasma NPTX1 levels (β = 0.261, p = 0.007), and this association remained significant after FDR correction across the seven vaccination-profile analyses (q = 0.049). Vaccination-profile breadth was not associated with plasma Aβ40 (β = 0.007, p = 0.948), and no significant associations were observed with the remaining plasma biomarkers. These findings provide preliminary evidence of an association between vaccination-profile breadth and plasma NPTX1, although this result should be interpreted cautiously given the exploratory nature of the analyses.

### CSF Replication Analyses

A set of 12 CSF analyses were performed to evaluate vaccination-related associations with the available CSF biomarkers (Supplementary Table S4). Overall, the plasma findings were not consistently reflected in the CSF analyses. However, herpes zoster vaccination was nominally associated with lower CSF Aβ42 (β = −0.646, p = 0.024) and lower CSF pTau (β = −0.701, p = 0.044). Vaccination-profile breadth was not associated with CSF Aβ42 (β = −0.153, p = 0.216) or CSF pTau (β = −0.039, p = 0.789). None of the CSF associations remained significant after FDR correction across the complete CSF analysis family. Sample sizes were smaller and varied across models (N = 53–66). Overall, the CSF analyses provided limited support for the exploratory plasma findings. Although herpes zoster vaccination showed nominal associations with lower CSF Aβ42 and pTau, these associations did not remain significant after correction. These results do not provide robust evidence that individual vaccine receipt or vaccination-profile breadth was associated with the selected CSF Alzheimer’s disease biomarkers in this cohort.

### Viral History x Vaccination Profile Breadth Interactions

A set of interaction analyses was conducted to explore whether viral exposure modified the association between vaccination-profile breadth and plasma Alzheimer’s disease-related biomarkers (Supplementary Table S5). 21 adjusted models examined interactions involving COVID-19, herpes zoster, or varicella history across the seven plasma biomarkers. None of the interaction terms reached the conventional significance threshold, and none remained significant after FDR correction. These analyses therefore did not support the hypothesis that the association between vaccination-profile breadth and plasma Alzheimer’s disease-related biomarkers varied according to viral exposure. No significant effect modification was observed for COVID-19, herpes zoster, or varicella history in this cohort.

### Psychological Symptoms Analyses

Fourteen adjusted regression models were conducted to explore associations between psychological symptoms and plasma biomarkers (Supplementary Table S6). Anxiety symptoms were assessed using the GAI, whereas depressive symptoms were assessed using the GDS. A small positive association was observed between depressive symptoms and plasma pTau217 (β = 0.269, p = 0.007), although this association did not remain significant after FDR correction (q = 0.098). No significant associations were observed between GAI scores and any of the plasma biomarkers. These findings provide preliminary evidence of an association between greater depressive symptoms and higher plasma pTau217, although this result should be interpreted cautiously given the exploratory nature of the analyses and the temporal separation between baseline plasma collection and the most recent GDS assessment.

### Associations across the full plasma NULISA panel

Across the full 115-analyte plasma NULISA panel, no individual vaccine-biomarker association survived FDR correction within the full NULISA panel. Several nominal associations were observed.

Pneumococcal vaccination was nominally associated with higher CALB2 and lower CD63, BACE1, DDC and ENO2. Influenza vaccination was nominally associated with higher VEGFA, CALB2, CHIT1, CXCL1 and Aβ40. Zoster/shingles vaccination was nominally associated with lower CCL11, FABP3 and SLIT2. Hepatitis A and B vaccination showed overlapping nominal associations with lower tau-related markers, NPY and POSTN, and higher PARK7. Because these associations did not survive correction across the full NULISA panel, they should be considered candidate signals for replication rather than definitive biomarker effects.

### Associations across the Olink CSF biomarker panel

The Olink candidate panel was available for a much smaller subset of participants (approximately 41 with measured CSF Olink values). No Olink biomarker was significantly associated with vaccination after adjustment or FDR correction, and no zoster/shingles-Olink association reached nominal P < 0.05. For the zoster comparison, several Olink markers showed numerically higher adjusted levels among vaccinated participants, including NTRK2, CD200, CDH6 and RGMB, but these did not approach corrected significance. This pattern is compatible with insufficient power in the small Olink subset and should not be read as evidence against a biological relationship.

## DISCUSSION

In this exploratory PREVENT-AD analysis, adult vaccination history was examined in relation to plasma AD biomarkers and inflammatory/proteomic measures from both CSF and plasma samples in a cohort enriched for preclinical AD risk. The analytic sample consisted primarily of cognitively normal older adults, consistent with the PREVENT-AD design as a longitudinal study of cognitively unimpaired individuals with a first-degree family history of sporadic Alzheimer-like dementia and repeated cognitive, neuroimaging, and biofluid assessments. This setting is particularly relevant because vaccination may influence AD risk most plausibly during the asymptomatic or early biological phase, before amyloid, tau, neuroinflammatory, and neurodegenerative cascades become self-sustaining.

Overall, the results do not support a broad, robust association between routine adult vaccination and lower plasma amyloid or tau pathology in this cross-sectional/proximal asymptomatic dataset. Influenza, zoster/shingles, pneumococcal, and Td/Tdap vaccination were not associated with FDR-significant differences in the principal AD biomarker panel examining tau and amyloid markers. COVID vaccination could not be meaningfully evaluated because nearly all participants were vaccinated. Nevertheless, the association between a wider vaccination profile breadth and a higher NPTX1 level remained significant after FDR correction. The analyses also revealed that greater depressive symptoms were associated with greater pTau217 plasma level, yet this association did not remain significant after FDR correction. Taken together, although epidemiological studies increasingly suggest that several vaccines are associated with lower dementia incidence, the present PREVENT-AD biomarker analysis does not yet demonstrate a clear plasma AD-biomarker signature that would explain such protection. The association between a broader vaccination profile breadth and a higher level might represent a distinct synaptic-related signal and would require further investigation. Furthermore, the plasma findings were not replicated in the CSF.

The strongest AD-biomarker signal was observed for hepatitis B vaccination, which was associated with lower plasma total tau/MAPT, p-tau181, and p-tau231 after correction within the AD biomarker panel. This is biologically intriguing because tau-related plasma markers are increasingly viewed as sensitive indicators of AD-relevant neuronal stress and early tau pathology. Nevertheless, one should approach this observation with caution. Vaccination records for Hepatitis A and B showed a strong connection, possibly indicating a distinct group with unique backgrounds (job, travel, finances, healthcare access, health habits or employment in the health care system) instead of a direct biological response to the vaccines themselves. At this stage, the hepatitis B finding should be framed as hypothesis-generating rather than evidence that hepatitis B vaccination directly modifies tau biology.

An intriguing potential interpretation of the hepatitis vaccination-associated tau signature arises from this antimicrobial-defense framework. Emerging evidence suggests that phosphorylated tau, like amyloid-β, may participate in host responses to viral infection. Viral infection can induce tau hyperphosphorylation, and phosphorylated tau has been reported to interact with viral particles and reduce infectivity. Under such a model, lower plasma tau and p-tau levels among vaccinated individuals could theoretically reflect a reduced cumulative requirement for infection-associated neuronal innate immune responses rather than direct suppression of tau production by vaccination. However, the present findings do not establish such a mechanism. Hepatitis A and B viruses are predominantly hepatotropic rather than neurotropic, vaccination timing and infection burden were not modeled, and hepatitis A and B vaccination were highly correlated in this cohort. The observed associations may therefore also reflect systemic inflammatory effects or unmeasured behavioral, occupational, socioeconomic, or healthcare-engagement characteristics associated with hepatitis vaccination.

The broader NULISA and Olink analyses also support a conservative interpretation. Across the complete NULISA panel, no biomarker association survived correction across the full set of analytes, although several nominal signals may help generate candidate immune pathways for future study. The Olink CSF analysis faced significant limitations due to a small number of volunteers who underwent lumbar puncture. It did not reveal nominal or FDR-significant vaccine-associated changes. Together, these findings suggest that any vaccine-related biological effects in this cohort are likely to be modest, heterogeneous, dependent on vaccine class, timing, host immune state, or genotype, and not easily captured by a single cross-sectional plasma profile.

These data remain valuable because they begin to bridge a major gap between population-level vaccine epidemiology and molecular AD biology. The questionnaire captured several adult vaccinations, including influenza, zoster/shingles, pneumococcal, Td/Tdap, VRS, COVID-19, hepatitis A, hepatitis B, and RSV, as well as prior viral infection history including varicella, shingles, recurrent herpes labialis, genital herpes, EBV, CMV, influenza, pneumonia, COVID-19, viral CNS infection, hepatitis, severe viral infection, and perceived post-infectious cognitive worsening. The present analyses also extended beyond individual vaccine exposures to examine vaccination profile breadth, selected viral-history, available CSF biomarkers, and psychological symptoms. Although these exploratory analyses yielded limited evidence after multiple-testing correction, they provide the foundation for future longitudinal studies incorporating vaccine timing, infection burden, herpesvirus reactivation history, and repeated biomarker measurements. This broader framework provides an important foundation for future models that examine not only individual vaccines, but also cumulative vaccination burden, infection burden, herpesvirus reactivation history, and their interaction with AD biomarkers.

### Limitations and future directions

The principal limitation is that the current analysis is cross-sectional at a baseline/proximal visit. Therefore, the results describe biomarker differences by vaccination status, not longitudinal biomarker change after vaccination. Vaccination exposure was also self-reported and coded mainly as binary history, without precise timing, number of doses, vaccine formulation, interval since vaccination, immune response magnitude, or antibody titers. This is especially important for zoster vaccination, where live and recombinant vaccines may differ biologically, and for COVID vaccination, where near-universal uptake prevented meaningful comparison. None of the participants involved in this study displayed long COVID symptoms; those who did eventually left the research program years ago. The Olink CSF subset was small, limiting power to detect immune-proteomic effects. A major challenge persists with residual confounding factors: vaccination status could be a proxy for health-seeking behaviours, healthcare access, education, travel, occupational exposures, comorbidities, and general preventive health attitudes.

Future analyses should prioritize longitudinal designs that align biomarker sampling with vaccination timing, distinguish vaccine formulation and dose series, and test cumulative vaccine exposure as well as infection burden. Models should adjust for age, sex, education, APOE4, baseline biomarker status, vascular risk, and healthcare-engagement proxies, and should examine interactions with APOE4 and sex given prior vaccine-dementia literature. The most informative next step would be to determine whether vaccination history predicts slower longitudinal change in plasma p-tau217/p-tau181, NfL, GFAP, inflammatory markers, amyloid/tau PET, or cognitive trajectories. Integration with infection history may also clarify whether apparent vaccine protection reflects prevention of pathogen reactivation, nonspecific immune remodeling, or broader host resilience.

## Data Availability

All data used in the present study are available upon reasonable request to the authors, subject to applicable ethical and privacy restrictions.

## Ethics approval and consent to participate

The PREVENT-AD study protocols, consent forms, and study procedures were approved by the McGill University Institutional Review Board and/or the Douglas Mental Health University Institute Research Ethics Board. All participants provided written informed consent.

## CRediT Authorship contribution statement

**Sophie Charland:** Conceptualization, Methodology, Investigation, Formal analysis, Writing – original draft, Writing – review & editing. **Mélissa Savard:** Formal analysis, Data curation. **Cynthia Picard**: Review & editing – visualization – data curation. **Judes Poirier:** Conceptualization, Methodology, Formal analysis, Data curation, Resources, Writing – original draft, Writing – review & editing, Supervision, Project administration, Funding acquisition.

## Acknowledgements

The authors would like to thank Laurence Maligne-Bruneau for her contribution to the collection of CSF and plasma samples.

## Data Availability

Data will be made available on request

