## Supplementary Material for "Adult Vaccination History and Plasma Biomarker Signatures in the Asymptomatic PREVENT-AD Cohort"

**Supplementary Table S1.** *Strongest co-vaccination correlations.*

| Vaccine 1 | Vaccine 2 | Phi | Complete pairs n |
|---|---|---|---|
| Hepatitis A | Hepatitis B | 0.8683 | 153 |
| Td/Tdap | Hepatitis B | 0.4547 | 138 |
| Td/Tdap | Hepatitis A | 0.3998 | 139 |
| Influenza | Pneumococcal | 0.3586 | 184 |
| Zoster/shingles | Hepatitis A | 0.3128 | 157 |

**Supplementary Table S2.** *Individual vaccine associations with plasma biomarkers*

| Exposure | Biomarker | Effect tested | B | SE | 95% CI for B | Standardized β | p | N | Adjusted $R^2$ | Partial $\eta^2$ | Model df | Robust SE | Global FDR q | Family FDR q | Analysis FDR q |
|---|---|---|---|---|---|---|---|---|---|---|---|---|---|---|---|
| Flu | Ratio Aβ42/Aβ40 | Flu vaccine | 0.017 | 0.016 | [-0.014, 0.048] | 0.274 | .269 | 135 | 0.065 | 0.009 | 5 | No | .736 | .570 | .503 |
| Flu | Aβ40 | Flu vaccine | 0.233 | 0.132 | [-0.029, 0.494] | 0.427 | .081 | 139 | 0.025 | 0.023 | 5 | No | .582 | .389 | .354 |
| Flu | Aβ42 | Flu vaccine | 0.234 | 0.183 | [-0.128, 0.597] | 0.313 | .203 | 139 | 0.012 | 0.012 | 5 | No | .693 | .541 | .474 |
| Flu | pTau217 | Flu vaccine | -0.294 | 0.132 | [-0.556, -0.032] | -0.574 | .028 | 107 | 0.105 | 0.047 | 5 | No | .434 | .233 | .210 |
| Flu | pTau181 | Flu vaccine | -0.083 | 0.133 | [-0.346, 0.179] | -0.146 | .530 | 139 | 0.110 | 0.003 | 5 | No | .874 | .828 | .805 |
| Flu | GFAP | Flu vaccine | 0.083 | 0.135 | [-0.183, 0.350] | 0.147 | .536 | 139 | 0.069 | 0.003 | 5 | No | .874 | .828 | .805 |
| Flu | NPTX1 | Flu vaccine | -0.047 | 0.104 | [-0.252, 0.158] | -0.109 | .653 | 139 | 0.032 | 0.002 | 5 | No | .928 | .867 | .857 |
| Pneumococcal | Ratio Aβ42/Aβ40 | Pneumococcal vaccine | -0.016 | 0.011 | [-0.037, 0.005] | -0.260 | .134 | 135 | 0.079 | 0.018 | 4 | Yes | .630 | .474 | .405 |
| Pneumococcal | Aβ40 | Pneumococcal vaccine | -0.247 | 0.092 | [-0.429, -0.065] | -0.454 | .008 | 139 | 0.042 | 0.051 | 4 | No | .242 | .176 | .193 |
| Pneumococcal | Aβ42 | Pneumococcal vaccine | -0.277 | 0.127 | [-0.527, -0.027] | -0.370 | .030 | 139 | 0.040 | 0.035 | 4 | No | .434 | .233 | .210 |
| Pneumococcal | pTau217 | Pneumococcal vaccine | 0.017 | 0.096 | [-0.173, 0.207] | 0.034 | .857 | 107 | 0.068 | 0.000 | 4 | No | .972 | .963 | .967 |
| Pneumococcal | pTau181 | Pneumococcal vaccine | -0.102 | 0.092 | [-0.285, 0.081] | -0.178 | .273 | 139 | 0.119 | 0.009 | 4 | No | .736 | .570 | .503 |

| Exposure | Biomarker | Effect tested | B | SE | 95% CI for B | Standardized β | p | N | Adjusted R² | Partial η² | Model df | Robust SE | Global FDR q | Family FDR q | Analysis FDR q |
|---|---|---|---|---|---|---|---|---|---|---|---|---|---|---|---|
| Pneumococcal | GFAP | Pneumococcal vaccine | -0.024 | 0.095 | [-0.212, 0.164] | -0.042 | .801 | 139 | 0.055 | 0.000 | 4 | No | .960 | .954 | .934 |
| Pneumococcal | NPTX1 | Pneumococcal vaccine | 0.161 | 0.071 | [0.020, 0.301] | 0.376 | .025 | 139 | 0.072 | 0.037 | 4 | No | .434 | .233 | .210 |
| TDC/Tdap | Ratio Aβ42/Aβ40 | TDC/Tdap vaccine | 0.040 | 0.016 | [0.009, 0.072] | 0.638 | .011 | 109 | 0.087 | 0.060 | 4 | No | .277 | .176 | .193 |
| TDC/Tdap | Aβ40 | TDC/Tdap vaccine | 0.147 | 0.142 | [-0.134, 0.428] | 0.263 | .301 | 114 | 0.007 | 0.010 | 4 | No | .745 | .602 | .527 |
| TDC/Tdap | Aβ42 | TDC/Tdap vaccine | -0.002 | 0.189 | [-0.376, 0.372] | -0.002 | .993 | 114 | 0.000 | 0.000 | 4 | No | 1.000 | .993 | .993 |
| TDC/Tdap | pTau217 | TDC/Tdap vaccine | -0.086 | 0.143 | [-0.371, 0.200] | -0.166 | .552 | 89 | 0.083 | 0.004 | 4 | No | .874 | .828 | .805 |
| TDC/Tdap | pTau181 | TDC/Tdap vaccine | -0.072 | 0.132 | [-0.333, 0.189] | -0.131 | .585 | 114 | 0.115 | 0.003 | 4 | No | .886 | .851 | .819 |
| TDC/Tdap | GFAP | TDC/Tdap vaccine | 0.158 | 0.136 | [-0.111, 0.428] | 0.284 | .247 | 114 | 0.077 | 0.012 | 4 | No | .718 | .570 | .503 |
| TDC/Tdap | NPTX1 | TDC/Tdap vaccine | -0.145 | 0.104 | [-0.352, 0.062] | -0.334 | .167 | 114 | 0.110 | 0.017 | 4 | No | .630 | .501 | .450 |
| VRS/RSV | Ratio Aβ42/Aβ40 | VRS/RSV vaccine | -0.002 | 0.017 | [-0.036, 0.031] | -0.038 | .884 | 113 | 0.072 | 0.000 | 4 | No | .972 | .963 | .967 |
| VRS/RSV | Aβ40 | VRS/RSV vaccine | -0.216 | 0.145 | [-0.505, 0.072] | -0.394 | .139 | 116 | 0.024 | 0.020 | 4 | No | .630 | .474 | .405 |
| VRS/RSV | Aβ42 | VRS/RSV vaccine | -0.378 | 0.195 | [-0.764, 0.008] | -0.503 | .055 | 116 | 0.062 | 0.033 | 4 | No | .461 | .293 | .275 |
| VRS/RSV | pTau217 | VRS/RSV vaccine | -0.177 | 0.151 | [-0.477, 0.123] | -0.368 | .245 | 91 | 0.048 | 0.016 | 4 | No | .718 | .570 | .503 |

| Exposure | Biomarker | Effect tested | B | SE | 95% CI for B | Standardized β | p | N | Adjusted $R^2$ | Partial $\eta^2$ | Model df | Robust SE | Global FDR q | Family FDR q | Analysis FDR q |
|---|---|---|---|---|---|---|---|---|---|---|---|---|---|---|---|
| VRS/RSV | pTau181 | VRS/RSV vaccine | -0.130 | 0.132 | [-0.391, 0.131] | -0.249 | .326 | 116 | 0.109 | 0.009 | 4 | No | .769 | .626 | .543 |
| VRS/RSV | GFAP | VRS/RSV vaccine | 0.059 | 0.154 | [-0.247, 0.365] | 0.100 | .702 | 116 | 0.056 | 0.001 | 4 | No | .935 | .887 | .877 |
| VRS/RSV | NPTX1 | VRS/RSV vaccine | 0.007 | 0.110 | [-0.212, 0.225] | 0.016 | .952 | 116 | 0.042 | 0.000 | 4 | No | .978 | .972 | .980 |
| Herpes zoster | Ratio Aβ42/Aβ40 | Herpes zoster vaccine | -0.031 | 0.015 | [-0.061, -0.001] | -0.491 | .041 | 135 | 0.092 | 0.032 | 4 | No | .434 | .246 | .239 |
| Herpes zoster | Aβ40 | Herpes zoster vaccine | 0.244 | 0.145 | [-0.043, 0.531] | 0.449 | .095 | 139 | 0.011 | 0.021 | 4 | No | .604 | .415 | .369 |
| Herpes zoster | Aβ42 | Herpes zoster vaccine | -0.012 | 0.200 | [-0.408, 0.384] | -0.016 | .951 | 139 | 0.005 | 0.000 | 4 | No | .978 | .972 | .980 |
| Herpes zoster | pTau217 | Herpes zoster vaccine | 0.219 | 0.134 | [-0.047, 0.485] | 0.427 | .106 | 107 | 0.092 | 0.025 | 4 | No | .604 | .424 | .371 |
| Herpes zoster | pTau181 | Herpes zoster vaccine | 0.063 | 0.144 | [-0.222, 0.349] | 0.111 | .661 | 139 | 0.113 | 0.001 | 4 | No | .928 | .867 | .857 |
| Herpes zoster | GFAP | Herpes zoster vaccine | 0.048 | 0.148 | [-0.245, 0.340] | 0.084 | .748 | 139 | 0.055 | 0.001 | 4 | No | .935 | .921 | .903 |
| Herpes zoster | NPTX1 | Herpes zoster vaccine | 0.145 | 0.112 | [-0.076, 0.367] | 0.340 | .197 | 139 | 0.049 | 0.012 | 4 | No | .693 | .541 | .474 |

**Supplementary Table S3.** *Vaccination Profile Breadth Associations with Plasma Biomarkers*

| Exposure | Biomarker | Effect tested | B | SE | 95% CI for B | Standardized β | p | N | Adjusted $R^2$ | Partial $\eta^2$ | Model df | Robust SE | Global FDR q | Family FDR q | Analysis FDR q |
|---|---|---|---|---|---|---|---|---|---|---|---|---|---|---|---|
| Vaccination-profile breadth | Ratio Aβ42/Aβ40 | Vaccination-profile breadth | -0.014 | 0.006 | [-0.026, -0.001] | -0.211 | .034 | 100 | 0.086 | 0.047 | 4 | No | .434 | .233 | .119 |
| Vaccination-profile breadth | Aβ40 | Vaccination-profile breadth | 0.004 | 0.057 | [-0.110, 0.117] | 0.007 | .948 | 103 | 0.000 | 0.000 | 4 | No | — | — | .948 |
| Vaccination-profile breadth | Aβ42 | Vaccination-profile breadth | -0.066 | 0.077 | [-0.219, 0.086] | -0.086 | .391 | 103 | 0.000 | 0.008 | 4 | No | .809 | .670 | .684 |
| Vaccination-profile breadth | pTau217 | Vaccination-profile breadth | 0.009 | 0.061 | [-0.112, 0.129] | 0.016 | .888 | 77 | 0.077 | 0.000 | 4 | No | .972 | .963 | .948 |
| Vaccination-profile breadth | pTau181 | Vaccination-profile breadth | -0.019 | 0.045 | [-0.108, 0.069] | -0.041 | .668 | 103 | 0.068 | 0.002 | 4 | No | .928 | .867 | .935 |
| Vaccination-profile breadth | GFAP | Vaccination-profile breadth | -0.058 | 0.063 | [-0.183, 0.068] | -0.098 | .382 | 103 | 0.049 | 0.010 | 4 | Yes | .809 | .670 | .684 |
| Vaccination-profile breadth | NPTX1 | Vaccination-profile breadth | 0.116 | 0.040 | [0.037, 0.196] | 0.261 | .007 | 103 | 0.123 | 0.074 | 4 | Yes | .242 | .176 | .049 |

**Supplementary Table S4.** *CSF Replication Analyses*

| Exposure | Biomarker | Effect tested | B | SE | 95% CI for B | Standardized β | p | N | Adjusted R² | Partial η² | Model df | Robust SE | Global FDR q | Family FDR q | Analysis FDR q |
|---|---|---|---|---|---|---|---|---|---|---|---|---|---|---|---|
| Individual vaccine | Aβ42 (CSF) | Flu vaccine | 123.959 | 97.145 | [-70.428, 318.346] | 0.363 | .207 | 66 | 0.288 | 0.027 | 6 | No | .693 | .655 | .432 |
| Individual vaccine | Aβ42 (CSF) | Pneumococcal vaccine | -163.630 | 84.539 | [-332.792, 5.533] | -0.480 | .058 | 66 | 0.288 | 0.060 | 6 | No | .461 | .377 | .232 |
| Individual vaccine | Aβ42 (CSF) | TDC/Tdap vaccine | 213.692 | 149.908 | [-87.719, 515.103] | 0.590 | .160 | 55 | 0.288 | 0.041 | 6 | No | .630 | .655 | .432 |
| Individual vaccine | Aβ42 (CSF) | VRS/RSV vaccine | -32.531 | 108.925 | [-251.785, 186.723] | -0.107 | .767 | 53 | 0.288 | 0.002 | 6 | No | .942 | .969 | .861 |
| Individual vaccine | Aβ42 (CSF) | Herpes zoster vaccine | -220.305 | 94.967 | [-410.269, -30.342] | -0.646 | .024 | 66 | 0.288 | 0.082 | 6 | No | .434 | .312 | .232 |
| Vaccination-profile breadth | Aβ42 (CSF) | Vaccination-profile breadth | -50.187 | 40.040 | [-130.784, 30.409] | -0.153 | .216 | 51 | 0.274 | 0.033 | 4 | No | — | — | .432 |
| Individual vaccine | pTau (CSF) | Vaccination-profile breadth | -0.753 | 2.796 | [-6.381, 4.876] | -0.039 | .789 | 51 | 0.000 | 0.002 | 4 | No | .953 | .969 | .861 |
| Individual vaccine | pTau (CSF) | Flu vaccine | 6.382 | 6.280 | [-6.185, 18.948] | 0.333 | .314 | 66 | 0.000 | 0.017 | 4 | No | .753 | .710 | .471 |

| Exposure | Biomarker | Effect tested | B | SE | 95% CI for B | Standardized β | p | N | Adjusted $R^2$ | Partial $\eta^2$ | Model df | Robust SE | Global FDR q | Family FDR q | Analysis FDR q |
|---|---|---|---|---|---|---|---|---|---|---|---|---|---|---|---|
| Individual vaccine | pTau (CSF) | Pneumococcal vaccine | 0.373 | 5.817 | [-11.267, 12.013] | 0.019 | .949 | 66 | 0.000 | 0.000 | 4 | No | .978 | .969 | .949 |
| Individual vaccine | pTau (CSF) | TDC/Tdap vaccine | 2.823 | 8.110 | [-13.467, 19.113] | 0.139 | .729 | 55 | 0.000 | 0.002 | 4 | No | .935 | .969 | .861 |
| Individual vaccine | pTau (CSF) | VRS/RSV vaccine | 8.003 | 6.900 | [-5.886, 21.891] | 0.480 | .252 | 53 | 0.000 | 0.028 | 4 | No | .718 | .655 | .432 |
| Individual vaccine | pTau (CSF) | Herpes zoster vaccine | -13.428 | 6.528 | [-26.481, -0.374] | -0.701 | .044 | 66 | 0.000 | 0.065 | 4 | No | .434 | .377 | .232 |

**Supplementary Table S5.** *Viral history x Vaccination Profile Breadth Interaction Analyses*

| Exposure | Biomarker | Effect tested | B | SE | 95% CI for B | Standardized β | p | N | Adjusted $R^2$ | Partial $\eta^2$ | Model df | Robust SE | Global FDR q | Family FDR q | Analysis FDR q |
|---|---|---|---|---|---|---|---|---|---|---|---|---|---|---|---|
| COVID | Ratio Aβ42/Aβ40 | COVID-19 × Vaccination-profile breadth | 0.010 | 0.016 | [-0.021, 0.041] | 0.151 | .536 | 97 | 0.082 | 0.004 | 6 | No | .874 | .871 | .889 |
| COVID | Aβ40 | COVID-19 × Vaccination-profile breadth | -0.158 | 0.128 | [-0.413, 0.097] | -0.304 | .221 | 99 | 0.000 | 0.016 | 6 | No | .695 | .870 | .773 |
| COVID | Aβ42 | COVID-19 × Vaccination-profile breadth | -0.121 | 0.175 | [-0.469, 0.227] | -0.168 | .492 | 99 | 0.000 | 0.005 | 6 | No | .874 | .871 | .889 |
| COVID | pTau217 | COVID-19 × Vaccination-profile breadth | -0.085 | 0.141 | [-0.367, 0.196] | -0.155 | .549 | 74 | 0.068 | 0.005 | 6 | No | .874 | .871 | .889 |
| COVID | pTau181 | COVID-19 × Vaccination-profile breadth | -0.221 | 0.138 | [-0.495, 0.053] | -0.369 | .112 | 99 | 0.105 | 0.027 | 6 | No | .604 | .752 | .701 |
| COVID | GFAP | COVID-19 × Vaccination-profile breadth | 0.020 | 0.140 | [-0.258, 0.298] | 0.034 | .888 | 99 | 0.035 | 0.000 | 6 | No | .972 | .948 | .932 |
| COVID | NPTX1 | COVID-19 × Vaccination-profile breadth | -0.047 | 0.098 | [-0.242, 0.149] | -0.107 | .635 | 99 | 0.150 | 0.002 | 6 | No | .928 | .909 | .889 |
| Varicella | Ratio Aβ42/Aβ40 | Vaccination-profile breadth × Varicella | 0.000 | 0.016 | [-0.031, 0.031] | 0.000 | 1.000 | 83 | 0.119 | 0.000 | 6 | Yes | 1.000 | 1.000 | 1.000 |
| Varicella | Aβ40 | Vaccination-profile breadth × Varicella | 0.189 | 0.184 | [-0.178, 0.555] | 0.328 | .309 | 84 | 0.000 | 0.013 | 6 | No | .753 | .871 | .725 |

| Exposure | Biomarker | Effect tested | B | SE | 95% CI for B | Standardized β | p | N | Adjusted $R^2$ | Partial $\eta^2$ | Model df | Robust SE | Global FDR q | Family FDR q | Analysis FDR q |
|---|---|---|---|---|---|---|---|---|---|---|---|---|---|---|---|
| Varicella | Aβ42 | Vaccination-profile breadth × Varicella | 0.225 | 0.258 | [-0.289, 0.738] | 0.280 | .386 | 84 | 0.000 | 0.010 | 6 | No | .809 | .871 | .725 |
| Varicella | pTau217 | Vaccination-profile breadth × Varicella | -0.178 | 0.243 | [-0.667, 0.310] | -0.332 | .587 | 60 | 0.027 | 0.010 | 6 | Yes | .886 | .880 | .725 |
| Varicella | pTau181 | Vaccination-profile breadth × Varicella | -0.127 | 0.092 | [-0.310, 0.056] | -0.226 | .243 | 84 | 0.065 | 0.007 | 6 | Yes | .718 | .871 | .725 |
| Varicella | GFAP | Vaccination-profile breadth × Varicella | 0.072 | 0.194 | [-0.314, 0.458] | 0.119 | .710 | 84 | 0.000 | 0.002 | 6 | No | .935 | .925 | .785 |
| Varicella | NPTX1 | Vaccination-profile breadth × Varicella | 0.074 | 0.095 | [-0.114, 0.262] | 0.172 | .524 | 84 | 0.110 | 0.004 | 6 | Yes | .874 | .871 | .725 |
| Herpes zoster | Ratio Aβ42/Aβ40 | Vaccination-profile breadth × Herpes zoster | 0.016 | 0.015 | [-0.014, 0.047] | 0.254 | .282 | 99 | 0.076 | 0.013 | 6 | No | .745 | .871 | .846 |
| Herpes zoster | Aβ40 | Vaccination-profile breadth × Herpes zoster | 0.126 | 0.140 | [-0.151, 0.403] | 0.223 | .370 | 102 | 0.000 | 0.008 | 6 | No | .809 | .871 | .946 |
| Herpes zoster | Aβ42 | Vaccination-profile breadth × Herpes zoster | 0.109 | 0.189 | [-0.265, 0.483] | 0.142 | .564 | 102 | 0.000 | 0.004 | 6 | No | .874 | .871 | .946 |
| Herpes zoster | pTau217 | Vaccination-profile breadth × Herpes zoster | 0.234 | 0.139 | [-0.043, 0.512] | 0.430 | .096 | 76 | 0.133 | 0.040 | 6 | No | .604 | .752 | .572 |

| Exposure | Biomarker | Effect tested | B | SE | 95% CI for B | Standardized β | p | N | Adjusted $R^2$ | Partial $\eta^2$ | Model df | Robust SE | Global FDR q | Family FDR q | Analysis FDR q |
|---|---|---|---|---|---|---|---|---|---|---|---|---|---|---|---|
| Herpes zoster | pTau181 | Vaccination-profile breadth × Herpes zoster | 0.224 | 0.138 | [-0.051, 0.499] | 0.371 | .109 | 102 | 0.108 | 0.027 | 6 | No | .604 | .752 | .572 |
| Herpes zoster | GFAP | Vaccination-profile breadth × Herpes zoster | 0.163 | 0.211 | [-0.256, 0.583] | 0.278 | .535 | 102 | 0.043 | 0.014 | 6 | Yes | .874 | .871 | .946 |
| Herpes zoster | NPTX1 | Vaccination-profile breadth × Herpes zoster | -0.063 | 0.102 | [-0.265, 0.140] | -0.140 | .542 | 102 | 0.110 | 0.004 | 6 | No | .874 | .871 | .946 |

**Supplementary Table S6.** *Psychological Symptoms Associated with Plasma Biomarkers*

| Exposure | Biomarker | Effect tested | B | SE | 95% CI for B | Standardized β | p | N | Adjusted $R^2$ | Partial $\eta^2$ | Model df | Robust SE | Global FDR q | Family FDR q | Analysis FDR q |
|---|---|---|---|---|---|---|---|---|---|---|---|---|---|---|---|
| GAI | Ratio Aβ42/Aβ40 | GAI score | -0.002 | 0.002 | [-0.006, 0.001] | -0.125 | .152 | 125 | 0.101 | 0.017 | 4 | No | .630 | .504 | .504 |
| GAI | Aβ40 | GAI score | -0.023 | 0.014 | [-0.051, 0.005] | -0.142 | .108 | 130 | 0.014 | 0.021 | 4 | No | .604 | .504 | .504 |
| GAI | Aβ42 | GAI score | -0.020 | 0.019 | [-0.058, 0.018] | -0.092 | .299 | 130 | 0.009 | 0.009 | 4 | No | .745 | .598 | .598 |
| GAI | pTau217 | GAI score | 0.018 | 0.014 | [-0.011, 0.046] | 0.122 | .216 | 98 | 0.082 | 0.016 | 4 | No | .694 | .504 | .504 |
| GAI | pTau181 | GAI score | -0.011 | 0.014 | [-0.039, 0.017] | -0.065 | .435 | 130 | 0.110 | 0.005 | 4 | No | .853 | .677 | .677 |
| GAI | GFAP | GAI score | 0.002 | 0.013 | [-0.024, 0.029] | 0.014 | .868 | 130 | 0.070 | 0.000 | 4 | No | .972 | .868 | .868 |
| GAI | NPTX1 | GAI score | 0.007 | 0.011 | [-0.014, 0.028] | 0.056 | .519 | 130 | 0.043 | 0.003 | 4 | No | .874 | .686 | .686 |
| GDS | Ratio Aβ42/Aβ40 | GDS score | -0.006 | 0.003 | [-0.012, 0.000] | -0.159 | .070 | 124 | 0.109 | 0.027 | 4 | No | .528 | .490 | .490 |
| GDS | Aβ40 | GDS score | -0.035 | 0.028 | [-0.091, 0.020] | -0.113 | .211 | 129 | 0.005 | 0.013 | 4 | No | .693 | .504 | .504 |
| GDS | Aβ42 | GDS score | -0.014 | 0.038 | [-0.090, 0.062] | -0.033 | .714 | 129 | 0.004 | 0.001 | 4 | No | .935 | .769 | .769 |
| GDS | pTau217 | GDS score | 0.074 | 0.027 | [0.021, 0.127] | 0.269 | .007 | 97 | 0.142 | 0.077 | 4 | No | .242 | .098 | .098 |
| GDS | pTau181 | GDS score | 0.017 | 0.028 | [-0.038, 0.072] | 0.052 | .539 | 129 | 0.109 | 0.003 | 4 | No | .874 | .686 | .686 |
| GDS | GFAP | GDS score | 0.021 | 0.027 | [-0.031, 0.074] | 0.070 | .424 | 129 | 0.074 | 0.005 | 4 | No | .853 | .677 | .677 |
| GDS | NPTX1 | GDS score | 0.009 | 0.021 | [-0.033, 0.052] | 0.039 | .662 | 129 | 0.039 | 0.002 | 4 | No | .928 | .769 | .769 |
